# Clinical characteristics and fecal–oral transmission potential of patients with COVID-19

**DOI:** 10.1101/2020.05.02.20089094

**Authors:** Saibin Wang, Junwei Tu, Yijun Sheng

**Affiliations:** Department of Respiratory Medicine, Jinhua Municipal Central Hospital, No. 365, East Renmin Road, Jinhua 321000, Zhejiang Province, China

**Author notes:** **Corresponding authors:** Saibin Wang, MD, Department of Respiratory Medicine, Jinhua Municipal Central Hospital, No. 365, East Renmin Road, Jinhua 321000, Zhejiang Province, China.

**Keywords:** COVID-19, SARS-CoV-2, nucleic acid, fecal–oral transmission

## Abstract

**Background:** A significant proportion of patients with COVID-19 generate negative pharyngeal swab viral nucleic acid test results but test positive using fecal samples. However, fecal–oral transmission of COVID-19 has not been established to date. The purpose of this study was to evaluate the duration of fecal swab positivity in COVID-19 patients after pharyngeal swab nucleic acid test turned negative and to explore its potential for fecal–oral transmission.

**Methods:** A retrospective analysis of clinical records, laboratory results, and chest computed tomography (CT) findings of 17 COVID-19 patients confirmed by laboratory tests from January 22 to February 7, 2020 at a tertiary hospital was performed. The potential of fecal–oral transmission was assessed by detecting the presence of SARS-CoV-2 nucleic acid in fecal swab samples.

**Results:** A total of 16 patients (94.1%) had fever; other symptoms included dry cough, dyspnea, nausea, diarrhea, sore throat, fatigue, and muscle pain. Three patients had decreased white blood cell counts, 7 had decreased lymphocyte numbers, and 7 had increased C-reactive protein levels. Fecal samples of 11 patients tested positive for SARS-CoV-2 nucleic acid, of whom the time for the fecal samples to become SARS-CoV-2 nucleic acid-negative was longer in 10 patients than that for pharyngeal swab samples, and only one case exhibited a shorter time for his fecal sample to become SARS-CoV-2 nucleic acid-negative compared to his pharyngeal swab sample. The remaining 6 patients were negative for SARS-CoV-2 nucleic acid in fecal samples.

**Conclusion:** In COVID-19 patients who tested positive for SARS-CoV-2 nucleic acid in both pharyngeal swab and fecal samples, the time for the fecal samples to become SARS-CoV-2 nucleic acid-negative was generally longer than that in pharyngeal swab samples. However, there is currently no evidence demonstrating that the virus can be transmitted through the fecal–oral route.

## 1. INTRODUCTION

Coronavirus Disease 2019 (COVID-19) was first reported in Wuhan City, Hubei Province, China in December 2019. Subsequently, the epidemic broke out locally and spread to the entire country and many other countries and regions [1–5]. COVID-19 caused by severe acute respiratory syndrome coronavirus 2 (SARS-CoV-2) is a highly contagious disease, and the WHO has announced its outbreak as a public health emergency [2]. COVID-19 is mainly transmitted through direct contact and droplet spread [1]. However, SAV-CoV-2 nucleic acid was frequently detected in fecal samples of COVID-19 patients [4, 6].

Currently, the time for fecal samples of COVID-19 patients to become SARS-CoV-2 nucleic acid-negative is unclear, and whether COVID-19 has the potential of fecal–oral transmission is unknown. The answers to these questions are critical to the development of patient quarantine observation window, duration of treatment, and clarification of transmission routes of COVID-19. Therefore, to enhance the prevention and control of COVID-19 in China and even globally, we retrospectively collected and analyzed the clinical data of 17 COVID-19 patients confirmed by laboratory tests. In this study, we describe the clinical characteristics of confirmed COVID-19 patients and analyze the fecal–oral transmission potential of COVID-19.

## 2. METHODS

### Study design and patient inclusion

We retrospectively included 17 confirmed COVID-19 patients admitted to Jinhua Municipal Central hospital (Jinhua, China) from January 22 to February 7, 2020. The diagnosis of COVID-19 was based on the Prevention and Control Plan of Novel Coronavirus Pneumonia (Trial version 7th) published by the National Health Committee of China and the National Administration of Traditional Chinese Medicine [1]. Respiratory pharyngeal swab samples were analyzed by qRT-PCR for viral nucleic acid, and all 17 patients tested positive for SARS-CoV-2. In the present study, we determined that pharynx swab SARS-CoV-2 nucleic acid-negative was defined as three consecutive negative tests of pharynx swab nucleic acid, with a minimum interval of 24 hours between each time; fecal virus nucleic acid-negative was defined as two consecutive fecal samples SARS-CoV-2 nucleic acid tests negative, with a minimum interval of 24 hours between the two. SARS-CoV-2 nucleic acid was tested using a kit recommended by the National Centers for Disease Control (BioGerm, Shanghai, China) according to the WHO guidelines for qRT-PCR [7]. All samples were tested simultaneously at the Centers for Disease Control of Jinhua city, China and the laboratory of Jinhua Municipal Central Hospital.

This study was reviewed and approved by the medical ethics committee of Jinhua Municipal Central Hospital (2020-llsc-172). The study data were collected anonymously, and written informed consent was waived by the ethics committee of the hospital for emerging infectious diseases..

### Data collection

We reviewed the clinical records, laboratory tests, and chest computed tomography (CT) findings of the 17 patients. All information was obtained through electronic medical records. Two researchers (SW and JT) independently reviewed data collection to confirm the accuracy of the data.

### Statistical analysis

Statistical analysis was performed using GraphPad Prism (ver. 7). Continuous variables were expressed as the mean and standard deviation. Categorical variables were represented by numbers (%). Between-group comparison, unpaired t-tests (normal distribution) or Kruskal-Wallis rank sum test (non-normal distribution), Pearson chi-squared tests or the Fisher’s exact was used, as appropriate. A P-value < 0.05 was considered as statistically significant.

## 3. RESULTS

All 17 patients had a history of epidemiological exposure, including 7 patients who came into contact with COVID-19 patients and 10 with a history of travel to Wuhan. Two patients had hypertension. The disease latency period was 1-11 days, with an average of 5.5 days. The clinical characteristics of the patients are shown in **Table 1**. Of the 17 patients, fever was the most common symptom (16/17), and all patients had low- to moderate-grade fever, with the highest temperature of 38.9°C. The second most common symptom (7/17) was dry cough, whereas sputum, headache, muscle pain, and gastrointestinal symptoms were rare.

A total of 14 patients had white blood cell counts within the normal range, and 3 patients had lower counts (<3.5×10^9/L); neutrophil count was within the normal range in 15 cases and lower (<1.8×10^^^9/L) in 2 cases; and lymphocyte count was within the normal range in 9 cases and lower (<1.1×10^9/L) in 7 cases. As shown in **Table 2**, there were no statistically significant of laboratory tests between the fecal virus nucleic acid-positive group and the fecal virus nucleic acid-negative group (P > 0.05).

**Table 1.** Baseline characteristics of the 17 COVID-19 patients

| Characteristics | n (%) |
| --- | --- |
| Male | 10 (58.8) |
| Age (year, Mean±SD) | 42 ± 17 |
| BMI (kg/m<sup>2</sup>, Mean±SD) | 24.0±3.0 |
| Smoking | 2 (11.8) |
| SBP (mmHg, Mean±SD) | 135.6±14.6 |
| DBP (mmHg, Mean±SD) | 87.0±12.2 |
| **Clinical symptoms** |  |
| Fever | 16 (94.1) |
| Dry cough | 7 (41.2) |
| Expectorant | 1 (5.9) |
| Sore throat | 2 (11.8) |
| Myalgia | 1 (5.9) |
| Shortness of breath | 4 (23.5) |
| Fatigue | 2 (11.8) |
| Nausea | 2 (11.8) |
| Diarrhea | 1 (5.9) |

COVID-19: corona virus disease 2019; BMI: body mass index.

**Table 2.** Laboratory tests and chest CT findings of the 17 COVID-19 patients

| Variables | Fecal samples positive for SARS-CoV-2 nucleic acid |  | P-value |
| --- | --- | --- | --- |
|  | Yes (n = 11) | No (n = 6) |  |
| Laboratory Tests |  |  |  |
| WBC (×10<sup>9</sup>/L) | 5.3 ± 1.5 | 4.9 ± 2.0 | 0.591 |
| Neutrophils (×10<sup>9</sup>/L) | 3.6 ± 1.3 | 2.8 ± 1.0 | 0.216 |
| Lymphocytes (×10<sup>9</sup>/L) | 1.3 ± 0.6 | 1.7 ± 1.0 | 0.401 |
| Hemoglobin (g/L) | 152.9 ± 17.7 | 140.3 ± 24.2 | 0.236 |
| platelets (×10<sup>9</sup>/L) | 191.3 ± 65.0 | 192.3 ± 58.0 | 0.974 |
| CRP (mg/L) | 13.8 ± 16.2 | 9.7 ± 4.4 | 0.557 |
| PT (S) | 12.7 ± 0.9 | 13.2 ± 1.6 | 0.395 |
| APTT (S) | 29.6 ± 2.3 | 30.5 ± 2.6 | 0.446 |
| D-Dimer (mg/L) | 0.3 ± 0.2 | 0.3 ± 0.1 | 0.889 |
| ALT (IU/L) | 34.0 ± 19.5 | 35.2 ± 16.5 | 0.902 |
| AST (IU/L) | 32.7 ± 11.6 | 38.0 ± 11.3 | 0.382 |
| TB (umol/L) | 17.3 ± 7.6 | 12.7 ± 5.3 | 0.206 |
| Creatinine (umol/L) | 73.8 ± 16.2 | 63.0 ± 11.6 | 0.172 |
| BUN (mmol/L) | 4.0 ± 0.5 | 3.3 ± 0.9 | 0.663 |
| CK (IU/L) | 105.5 ± 129.5 | 78.7 ± 29.1 | 0.628 |
| CK-MB (IU/L) | 9.6 ± 5.5 | 13.2 ± 4.0 | 0.186 |
| LDH (IU/L) | 457.5 ± 93.4 | 524.7 ± 174.2 | 0.311 |
| Lesion on CT scan |  |  | 0.938 |
| No lesion (%) | 2 (18.2) | 1 (16.7) |  |
| Bilateral lungs (%) | 9 (81.8) | 5 (83.3) |  |

WBC, white blood cell; CRP, C-reactive protein; PT, prothrombin time; APTT, activated partial thromboplastin time; ALT, alanine aminotransferase; AST, aspartate aminotransferase; TB, Total bilirubin; BUN, blood urea
nitrogen; CK, creatine kinase, CK-MB, creatine kinase-MB isoenzyme; LDH, lactic dehydrogenase

All 17 patients were treated with antiviral drugs (lopinavir/ritonavir and interferon-α2b), and five patients were treated with oxygen support (nasal catheter oxygen inhalation). A total of 14 patients were treated with empirical antibiotics and 4 patients were treated with hormones.

All 17 patients underwent chest CT examination. Fourteen patients showed typical chest CT findings for COVID-19 in bilateral lungs, i.e., multiple patchy ground-glass opacities in the lungs (**Figure 1**).

**Fig. 1.**
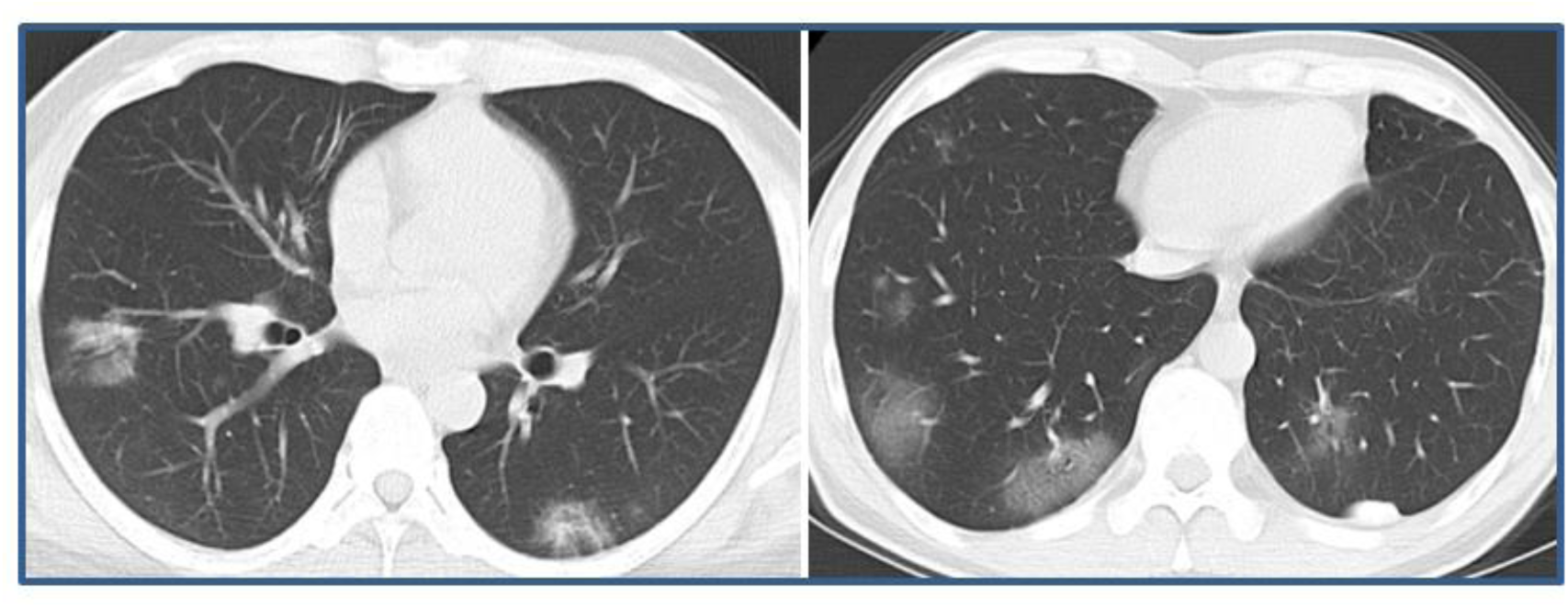
Transverse chest CT showing bilateral ground-glass opacity in COVID-19 patients.

Among the 17 patients, SARS-CoV-2 nucleic acid was detected in fecal samples of 11 (64.7%) patients, of whom the time for the fecal samples to become SARS-CoV-2 nucleic acid-negative in 10 patients (average time: 17 days, longest time: 40 days) was significantly longer than the pharyngeal swab samples, and only in one case exhibited an earlier time for the fecal sample to become SARS-CoV-2 nucleic acid-negative (8 days earlier) than the pharyngeal swab sample (**Figure 2**).

**Fig. 2.**
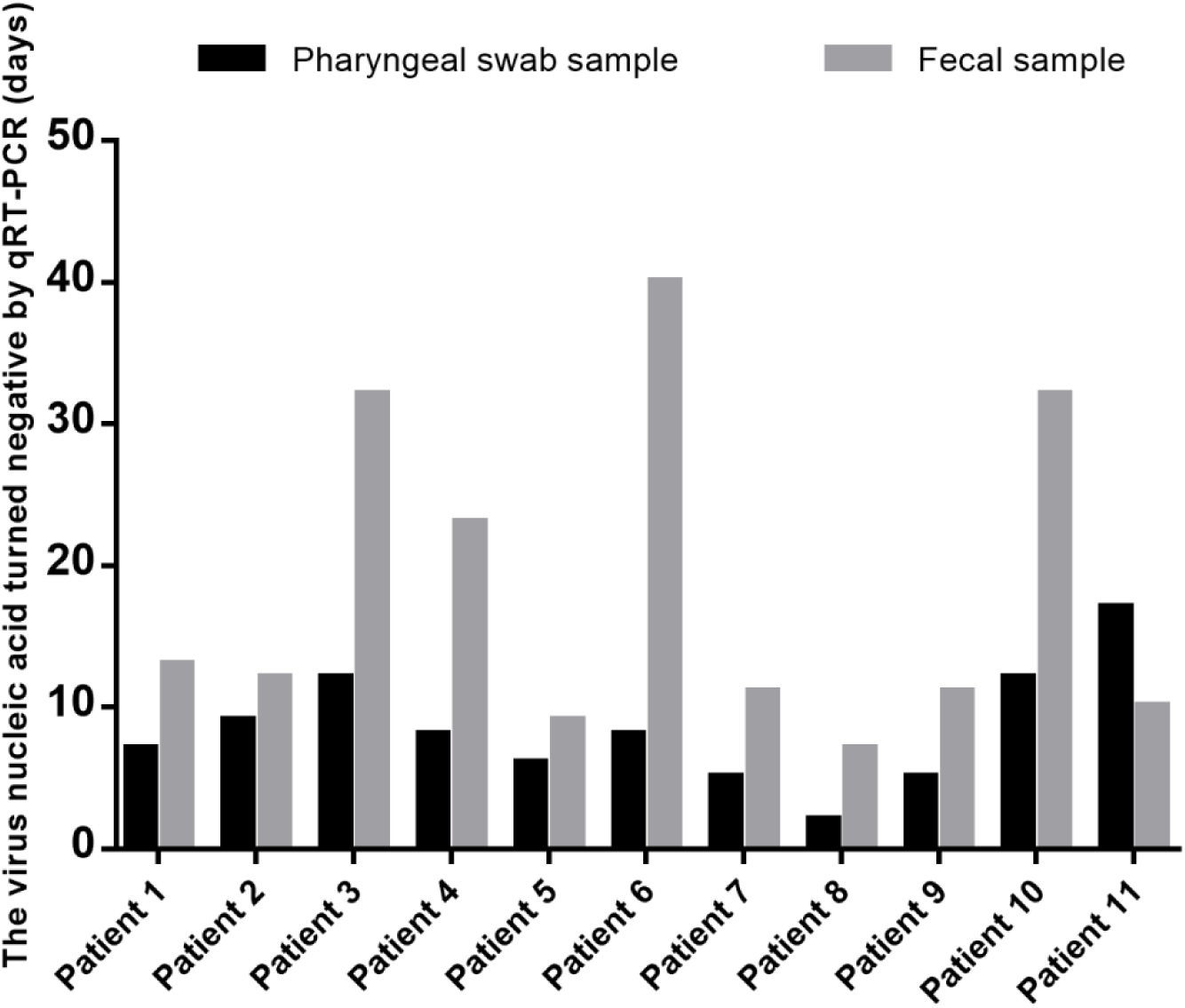
The duration of fecal samples and pharyngeal swab nucleic acid qRT-PCR test turned negative in 11 patients with COVID-19.

## 4. DISCUSSION

We report the clinical data of 17 COVID-19 patients confirmed by laboratory tests, including 2 severe and 15 regular cases, all of whom were cured and discharged. Among the 17 patients, 11 patients tested positive for SARS-CoV-2 nucleic acid using fecal specimens. Based on the investigation of our team, there is currently no evidence supporting the fecal–oral transmission in COVID-19 patients.

COVID-19 is highly contagious, and the population is generally susceptible to SARS-CoV-2 [1]. Based on the current epidemiology, it is believed that COVID-19 is mainly transmitted through direct contact and droplet spread. There is a possibility of aerosol transmission when people are exposed to high concentrations of aerosol for a long time in a relatively closed environment [1]. However, recently, it was found that the fecal samples of many COVID-19 patients were positive for SARS-CoV-2 nucleic acid [4, 6], which naturally raises the question: Can COVID-19 be transmitted through the fecal–oral route? SARS-CoV-2 is an enveloped, positive, single-stranded RNA coronavirus that is phylogenetically closely related to the SARS-CoV that caused the 2003 SARS outbreak (sequence similarity of up to 85%). SARS-CoV has been detected in fecal samples of SARS patients, and fecal samples have therefore become a choice of specimen examination for the early diagnosis of SARS [8–10]. Reportedly, the receptor (angiotensin-converting enzyme-II, ACE-II) of SARS-CoV-2 is highly expressed not only in type II alveolar epithelial cells, but also in esophageal and intestinal epithelial cells [11]. Moreover, the virus of SARS-CoV-2 has been successfully isolated from fecal samples of patients with COVID-19, suggesting the possibility of fecal–oral transmission of COVID-19 [12].

We found that the clinical manifestations of COVID-19 patients with positive feces and patients with negative feces were similar, including the common symptoms of fever and dry cough, and less common symptoms of muscle pain, general discomfort, and sore throat, and the severe symptom of dyspnea. Gastrointestinal symptoms such as nausea, vomiting, and diarrhea were rarely observed [7]. Based on observations of this group of patients, there was no significant correlation between digestive symptoms and fecal samples SARS-CoV-2-positive. Of note, it has been reported that in some atypical cases of COVID-19, gastrointestinal symptoms can be the primary complaint for patients [13].

In laboratory tests, COVID-19 patients might have lower lymphocyte counts and elevated CRP levels. However, not every patient exhibited these symptoms. Compared with laboratory tests, chest CT findings of COVID-19 patients were more consistent, i.e., 14 of 17 patients demonstrated typical lung changes, and thus CT has relatively high diagnostic value [14].

The main focus of this study was to investigate the possibility of fecal–oral transmission of COVID-19 infection. Of the 17 patients, 6 patients were from two families. Three patients of one family and 2 patients of another family tested positive for fecal SARS-CoV-2 nucleic acid, whereas another patient was negative. We reviewed the epidemiological history and living conditions of all patients. All of the patients lived in separate homes and maintained good hygiene habits before onset of disease. There was no evidence that they were infected with COVID-19 through the fecal–oral route rather than droplet spread or contact transmission. Further follow-up after hospital discharge also did not indicate that family members living with them in the same home were infected with COVID-19. However, the conclusion of no evidence of fecal–oral transmission in this study may be related to the small sample size and insufficient observation length for COVID-19. For example, previous studies have shown that SARS-CoV that causes SARS may be transmitted via the fecal–oral route[15].

This study is limited by the small sample size and retrospective research method. Several concerns should be considered when interpreting our results. First, there was still a risk for false negative results for PCR nucleic acid testing of pharyngeal swab and fecal samples. Second, some patients did not have fecal samples every day, and thus the time when SARS-CoV-2 nucleic acid results are negative may not be accurate in some patients. Third, there was no data of viral nucleic acid detection of lower respiratory tract specimens such as the SARS-CoV-2 nucleic acid detection of alveolar lavage fluid, and thus we cannot compare the time when SARS-CoV-2 nucleic acid detection results became negative in fecal and lower respiratory tract samples. In addition, we failed to explore the correlation between SARS-CoV-2 nucleic acid-positive results using fecal samples and disease severity, because only two severe cases were involved in this study.

In conclusion, based on the analytical results of this group of COVID-19 cases, the percentage of patients with fecal samples testing positive for SARS-CoV-2 nucleic acid was high, whereas the digestive symptoms of these patients were not significant. COVID-19 patients may have a variety of symptoms, mainly including fever and cough. We did not identify evidence for fecal–oral transmission of COVID-19.Considering the importance of this continuous global public health emergency, although the conclusions of this study are limited by the sample size, the results of this study may be used as reference for understanding the potential of fecal–oral transmission of COVID-19. Given the fact that the duration of SARS-CoV-2 nucleic acid-positive results using fecal samples is relatively long, sufficient attention should be paid to the disinfection and isolation during the recovery period of COVID-19 patients.

## Data Availability

The data can obtain from the corresponding author by request.

## Data Availability

The datasets used and/or analyzed during the current study are available from the corresponding author on reasonable request.

## Competing Interests

The authors declares that he has no competing interests.

## Author’ Contributions

S. W. contributed substantially to the study design, data analysis and interpretation, the writing of the manuscript, and takes responsibility for the integrity of the data and the accuracy of the data analysis. J. T. contributed to data collected, data analysis, data interpretation. Y. S. contributed to data interpretation and draft revision.

## Acknowledgements

We appreciate all the participants involved in the study. This study was supported by the Research Project of Emergency Prevention and Control of COVID-19 in Jinhua City (No. 2020XG-15).

## Notes

### Competing Interest Statement

The authors have declared no competing interest.

### Funding Statement

The study was supported by the science and technology project (2020XG-15)

